# Practical recommendations for staying physically active during the COVID-19 pandemic: A systematic literature review

**DOI:** 10.1101/2020.06.24.20138313

**Authors:** Ellen Bentlage, Achraf Ammar, Hamdi Chtourou, Khaled Trabelsi, Daniella How, Mona Ahmed, Michael Brach

## Abstract

**Background:** Due to home-confinement and social isolation of during the Covid-19 pandemic, reductions in performing physical activities were observed. A main consequence of inactivity is a poorer general health and a higher mortality rate. Therefore, it is important to inform the public about practical recommendations for staying physically active, especially during the current Covid-19 pandemic.

**Methods:** Through a systematic review of literature in two databases (Pubmed/Medline; Web of Science), studies were analysed which include practical recommendations for staying active during Covid-19 (Q1), or if they did not explicitly deal with Covid-19, research with useful results regarding the adaption to the present situation (Q2).

**Results:** Currently, there are 6 studies published which are related to the first research question. In total, 26 papers, were found to correlate to the second one.

**Conclusion:** Researchers need to be more specific in the exact recommendations for different age-groups and various health statuses. Key sources are the websites of the World Health Organization and the American Heart Association. In addition, exergames need to be adaptable to restrictions by product designers and to integrate social interaction-functions. Furthermore, it is essential that governmental actions need to be taken, with the help of researchers, to inform citizens about possible physical activities, with precise examples, clarification of benefits of the exercises, the exact exercise with duration, intensity and other substitute tasks for different age-groups and for people with diseases.

## Introduction

COVID-19, which first became public in December 2019, was declared a pandemic on the 12^th^ of March 2020 (WHO, 2020a), by which time the infectious disease had resulted in approximately 278,000 deaths worldwide (WHO, 2020b). Due to the rapid growth in the number of infections of Covid-19, lockdowns with home-confinement and social distancing were implemented (some still are implemented in many countries at time of authorship), in order to slow the spread of the virus, and to conserve treatment possibilities in public hospitals (Jiang et al., 2020). Practically this mean that gyms, public parks and schools were closed (Yarimkaya & Esenturk, 2020), among other facilities. This situation would negatively affect children and youth, who normally have an active lifestyle through active transport, school-related activities and sport participation (Hofman et al., 2019). Reduced physical activity as a result from social isolation measures is a serious concern particularly for older adults too, as they are typically more inactive compared to younger aged individuals and more prone to chronic diseases (Roschel et al., 2020). Results from a multicultural questionnaire, which was spread across Europe, Western Asia, North-Africa and to America, indicates that home confinement and social isolation measures could negatively affect all levels of physical activity (vigorous, moderate, walking and overall), along with increased daily sitting time by more than 28 percent (Ammar et al., 2020). A quantitative measurement of average steps for the duration of one week at the end of March 2020, assessed with the device Fitbit©, compared to the same period a year before, which analysed data from 30 million people, shows a fundamental reduction in average steps in almost all countries (Fitbit, Inc., 2020).

Research shows that physical inactivity is a high-risk factor for major disease morbidity (Hallal et al., 2012). A large-scale, prospective study has demonstrated when 30 min of day of sitting time is replaced by 30 minutes of day light-intensity PA a 24 percentage lower mortality-risk from cardiovascular causes (Dohrn et al., 2018). In summation, regular physical activity is healthy for the mind and the body (e.g. reducing high blood pressure, helping in managing weight, reducing risk of type 2 diabetes, stroke, heart disease, and various cancers), which are all variables that can increase susceptibility to the pandemic (WHO, 2020c). With some predictions estimating that the Covid-19 pandemic could possibly persist for 18 months (Ferguson et al., 2020), the negative impact on performing physical activity and increased sedentary behavior could persist with a serious detrimental affect on health. Therefore, practical recommendations for the performance of PA in times of a pandemic, such as Covid-19 are needed.

To our knowledge, no study to date has examined the possible relationship between quarantine/social isolation and the undertaking of deliberate physical activities within the home environment during these specialised conditions. Nevertheless, some tentative practical recommendations can be made based on the existing scholarly literature. Therefore, the purpose of this systematic review is to summarize the existing literature about possible practical recommendations for being physically active during this period of isolation.

In order to exploit scientific literature for this task, two research questions arise:

Q1. What has already been published in scientific journals regarding practical recommendations for physical activity during social distancing and home-confinement in the COVID-19 pandemic? Which recommendations or choices can be extracted?

Q2. Which topics and key factors for the performance of physical activities can be identified in journal articles for possible transfer to the present Covid-19 situation?

## Methods

The literature search was performed on May 16^th^, 2020 in the databases Pubmed/MEDLINE and Web of Science. There were no restrictions for study design, setting, country or time frame. For better quality, the MeSH (medical subject headings) thesaurus is used when possible.

We based the search strategy on the psychological construct “situation”, as defined by the component’s “person”, “environment” and “task”, which has been introduced for physical activity by Nitsch (2005). Brach et al (2017) applied this simple framework for describing how these components enable but also constrain a room for maneuver in individual mobility. Regarding the present topic, we didn’t apply search terms for the “person” component, because we didn’t have a special target group in mind. Regarding the “environment” component, we used four OR-combined terms. Three were MeSH Terms. The corresponding scope notes are:

- patient isolation (D010356): “*The segregation of patients with communicable or other diseases for a specified time. Isolation may be strict, in which movement and social contacts are limited; modified, where an effort to control specified aspects of care is made in order to prevent cross infection; or reverse, where the patient is secluded in a controlled or germ-free environment in order to protect him or her from cross infection*.”
- social isolation (D012934): “*The separation of individuals or groups resulting in the lack of or minimizing of social contact and/or communication. This separation may be accomplished by physical separation, by social barriers and by psychological mechanisms. In the latter, there may be interaction but no real communication*. “
- pandemics (D058873): “*Epidemics of infectious disease that have spread to many countries, often more than one continent, and usually affecting a large number of people*. “

Additionally, we added a fourth keyword for the research in the titles and abstracts, in order to find all articles related to confinement phases:

- “confinement”

The “task” component of the situation framework regards to physical activity. We referred to a search strategy from another review (Hinrichs & Brach 2012), who used the OR-combined MeSH terms “exercise”; “sports”; “physical activity”; physical fitness”; “motor activity”. In testing runs however, we missed papers which included physical activity, but not as a topic. Thus, we changed from MeSH terms to “search in title and abstract” and skipped “motor activity”.

The final search term for Pubmed was (“patient isolation”[MeSH Terms] OR “social isolation”[MeSH Terms] OR “confinement” [Title/Abstract] OR “pandemics”[MeSH Terms]) AND (“exercise”[Title/Abstract] OR “sports”[Title/Abstract] OR “physical activity”[Title/Abstract]).

The final search term for WebofScience (Topic search) was (“patient isolation” OR “social isolation” OR “pandemics” OR “confinement”) AND (“exercise” OR “sports” OR “physical activity” OR “physical fitness” OR “motor activity”).

We found 585 studies in PubMed/MEDLINE and 857 in the Web of Science (See Prisma Flow Diagram, figure 1). We uploaded them into the data management software Rayyan QCRI (Ouzzani et al., 2016) to analyze the data systematically. After removing 236 duplicates, there were 1.206 studies left. Two researchers (EB and MB) marked inclusion and exclusion independently. After this procedure, 998 studies were independently excluded and 179 conflicting marks were discussed and solved. Exclusion rules were animal and physiological studies, studies on diseases (e.g. autism) leading to social isolation, papers solely describing or analyzing the isolated situation, but without intervention.

**Figure 1:**
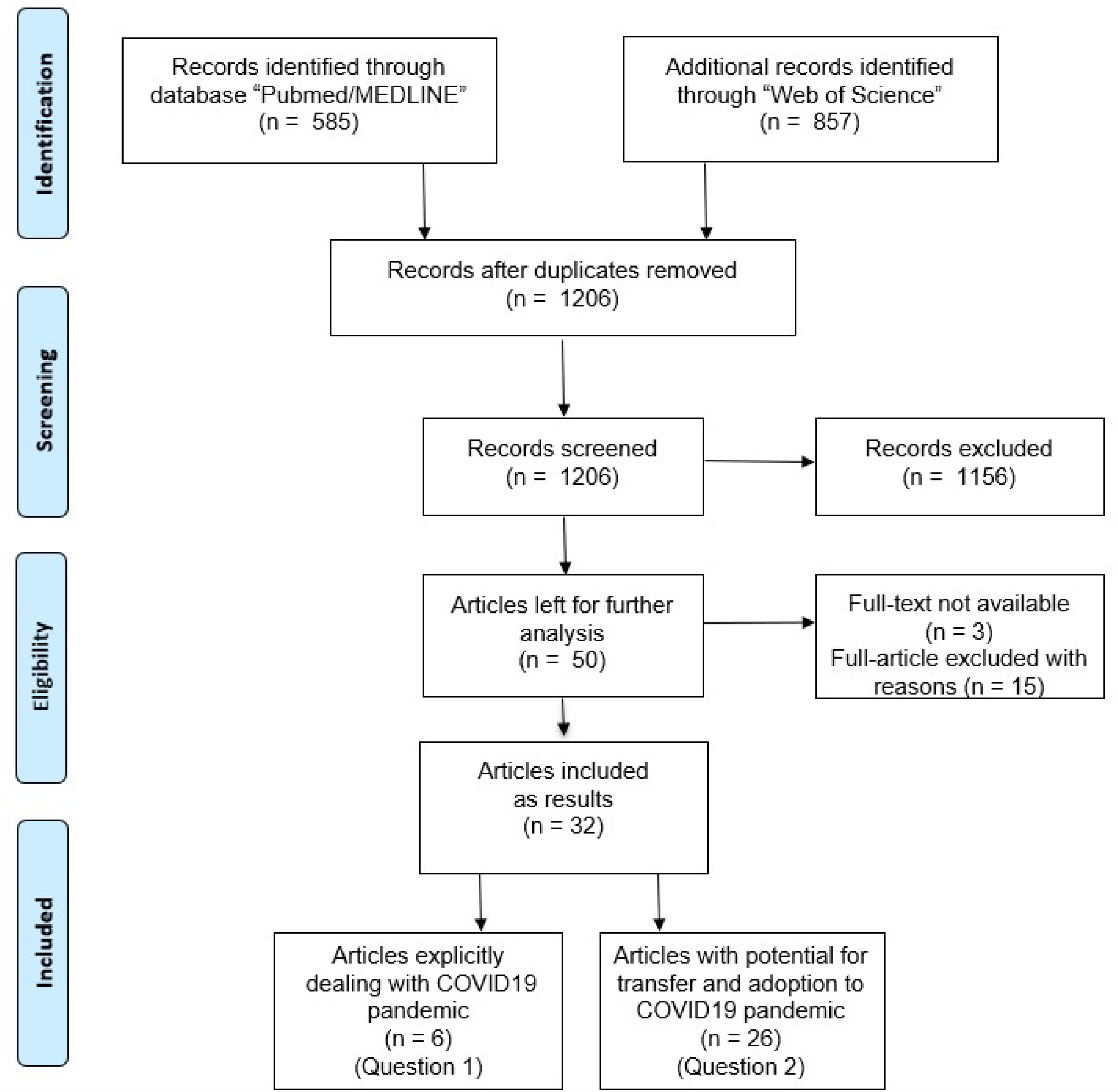
PRISMA flow chart.

After this screening procedure, 50 studies were left in the database. We found 47 full-texts, resulting in the other three abstract-only studies being excluded. After full-text scanning, a further 15 studies did not match with inclusion rules and were excluded. The remaining papers were six studies which explicitly dealt with physical activity (PA) in the Covid-19 pandemic (see table 1), and 26 articles which discussed PA-topics with the potential to adapt to the Covid-19 situation (see table 2).

**Table 1:**
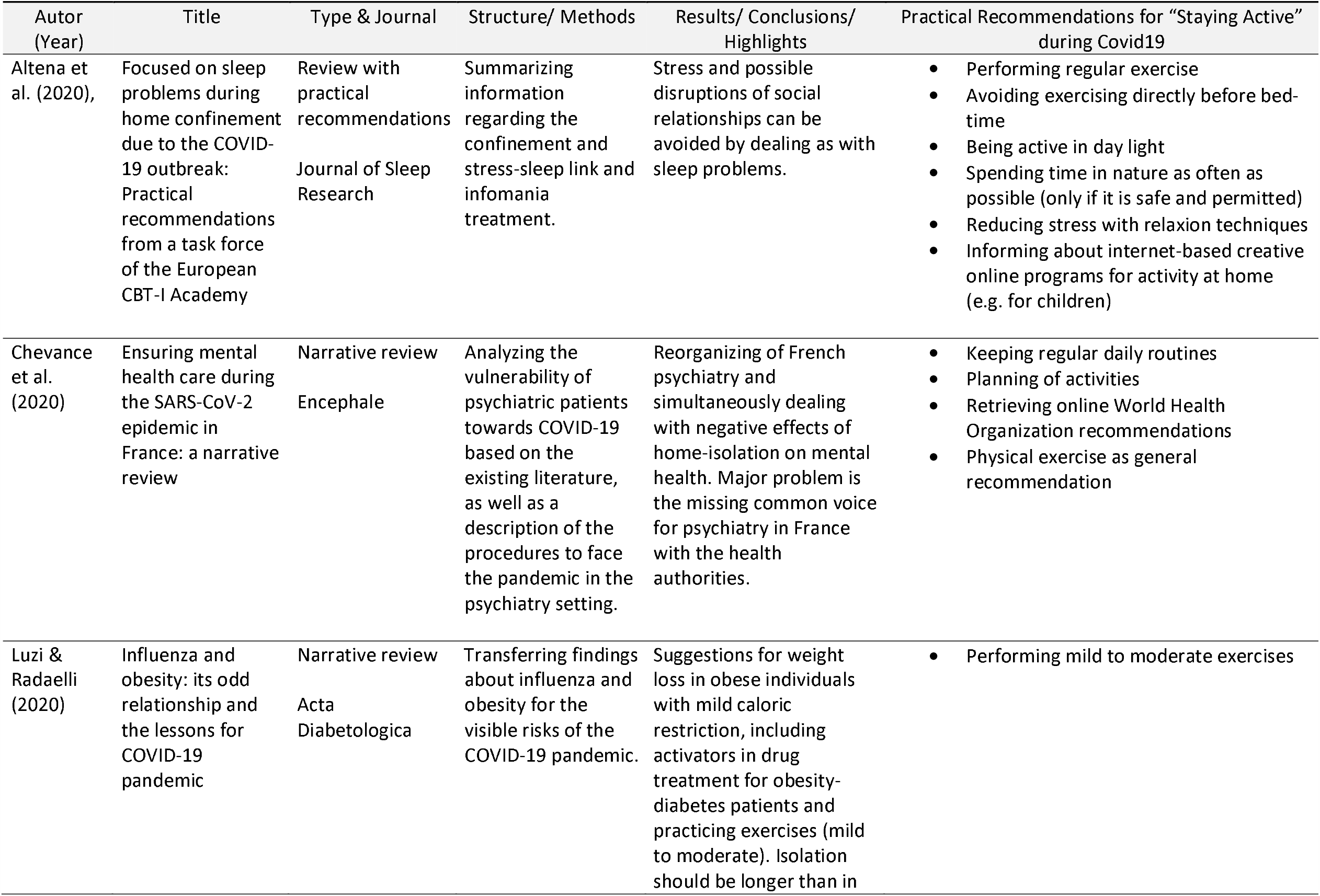
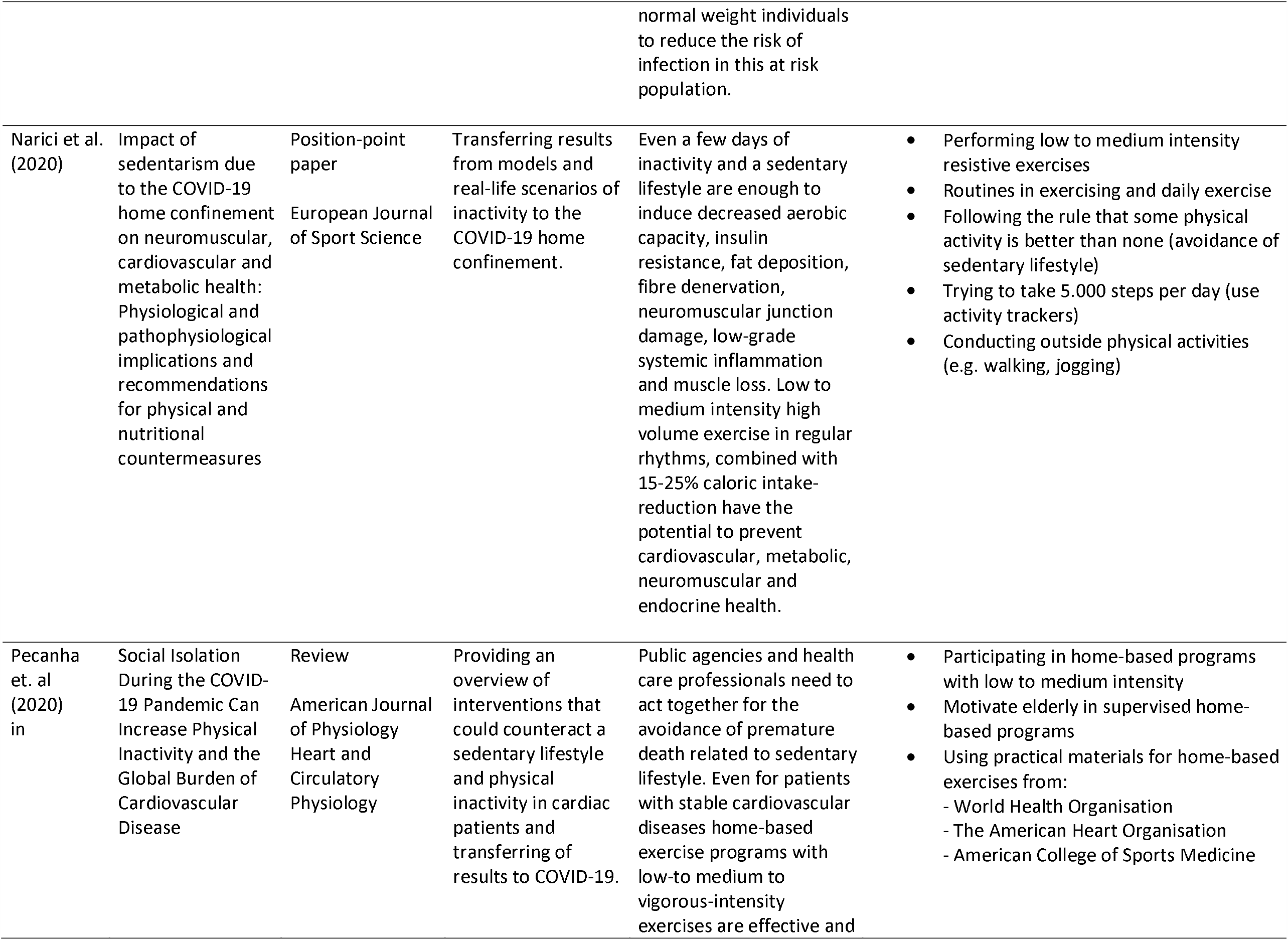
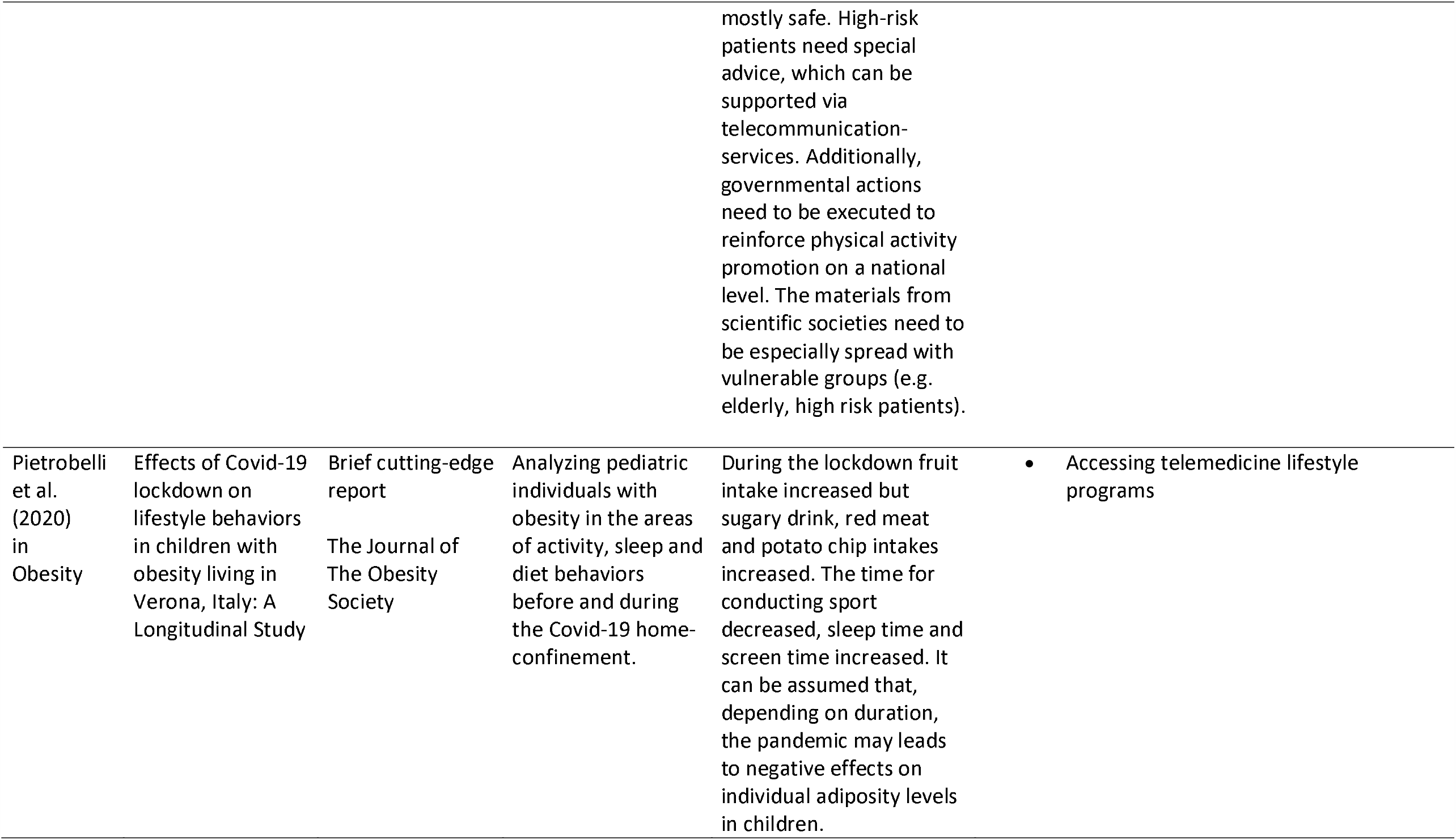
Practical Recommendations of studies for “Staying Active” during Covid-19

**Table 2:**
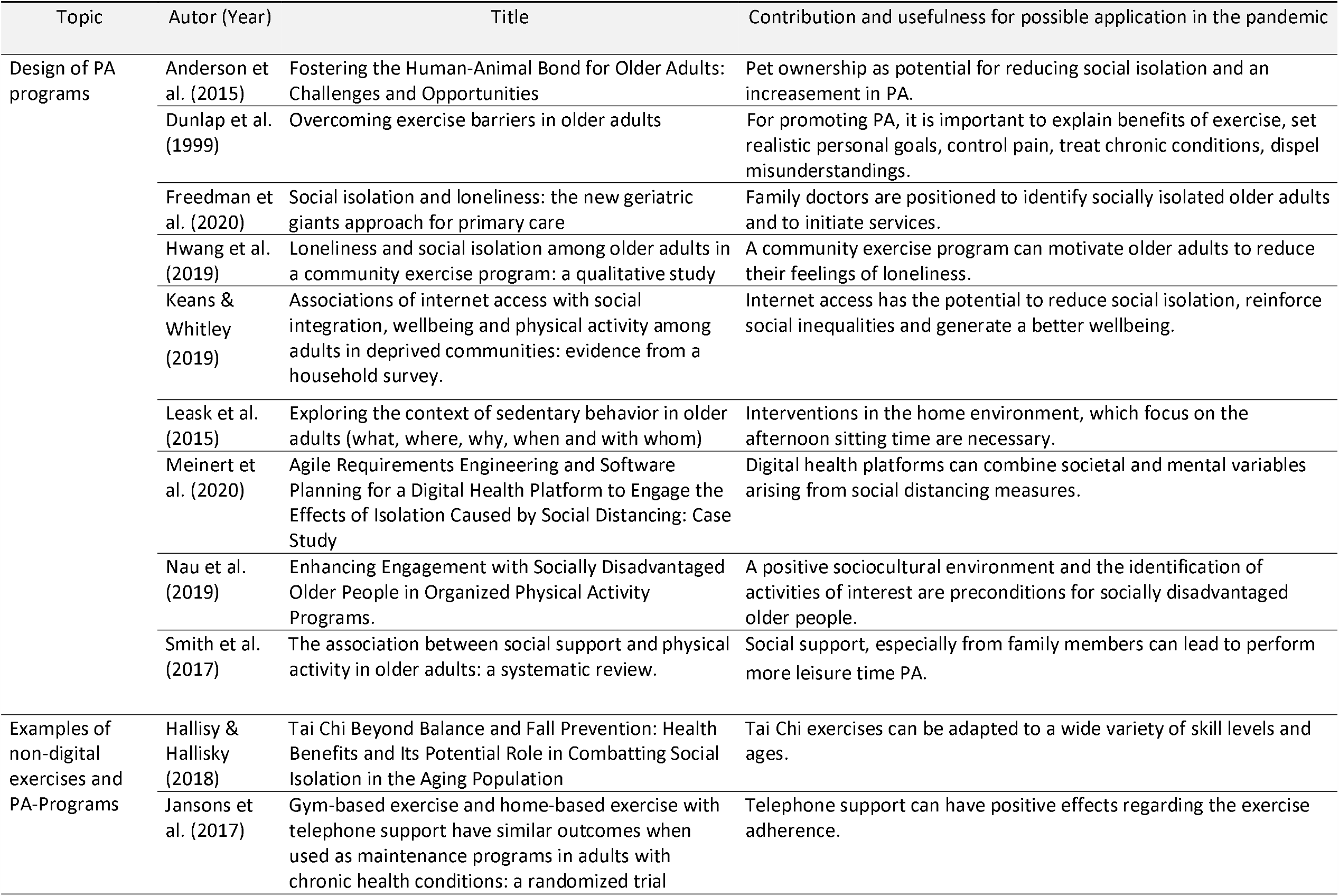
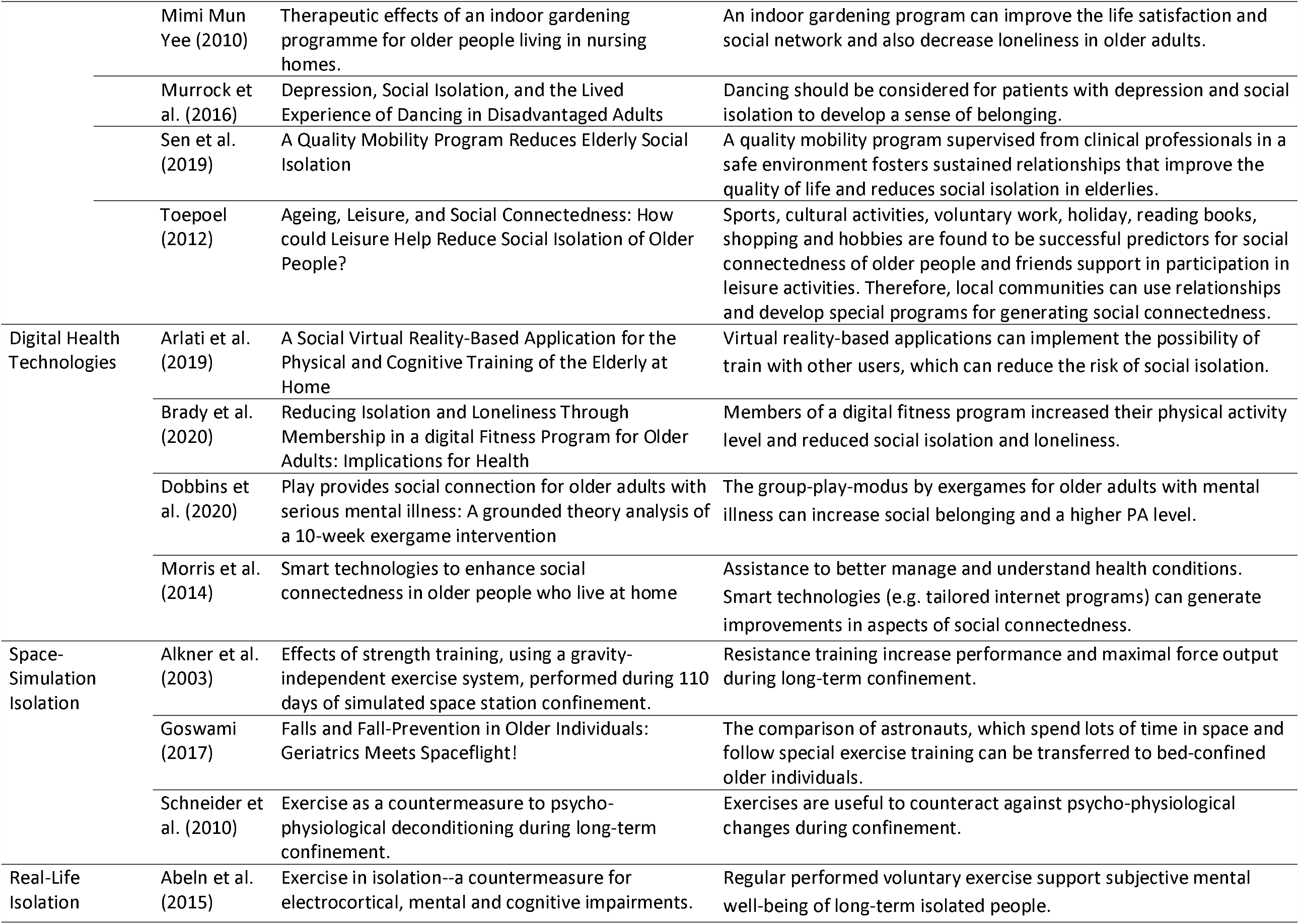
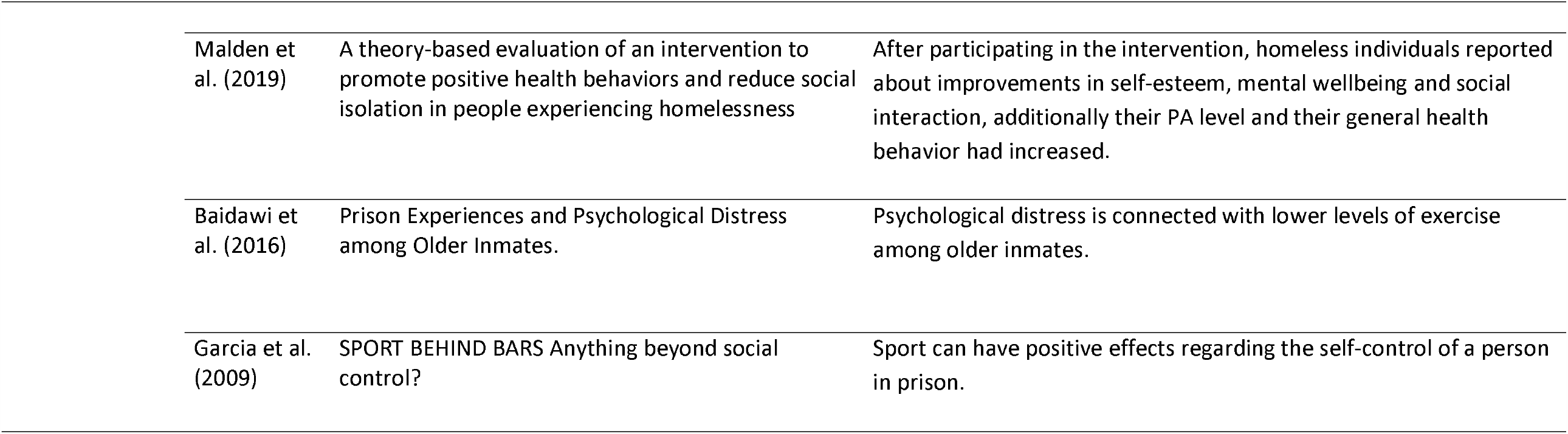
Useful results which could be adopted for the present situation

## Results

This systematic review gathered all data to practical recommendations for performing physical activity directly related to the Covid-19 restrictions and also to other isolation and confinement. Therefore, in the following paragraphs, we sketch the articles relating to question 1, practical recommendations for staying active during the Covid-19 pandemic, and summarise major points in table 1. After this procedure and with regard to the research question 2, we grouped the studies, which do not directly refer to the Covid-19 pandemic but could be adapted in relation to PA to the present situation into 5 topics (see table 2).

### Maintenance or increase of physical activity in the COVID-19 pandemic

Altena et al. (2020) largely describes the sleep problems during the current COVID-19 home confinement. Some aspects regarding the connection between sleep and physical activity are discussed, in particular, the effect of exercise on circadian rhythm. Low levels of activity during the day affect the sleep negatively. PA should be performed during the day, if possible in daylight and not immediately before bed-time. Major stress, because of the confinement in the current pandemic-situation, leads to sleep disruption. Physical exercise (e.g. mediation, muscle relaxion) can reduce the stress. Regular exercise is preventive and reduces the risk to increase the body weight.

The narrative review of Chevance et al. (2020) provides an overview of the mental health care in the Covid-19 epidemic in France where restrictions did not allow for physical exercise outside of the home and social interaction beyond the immediate household. Daily routine is essential and activities should be planned. Physical exercise belongs to the recommendations, but is not concretely described.

The article from Luzi & Radelli et al. (2020) analyses the relationship between obesity and influenza. It addresses the current COVID-19 pandemic and evidence from previous epidemics. The research established that one of the determinants in the severity of influenza viral infections in obese patients is the physical inactivity (sedentariness). PA and exercise are protective strategies to immunological and metabolic health. PA interventions showed potential for the modulation of inflammation, support of the immune system and improvement in vaccination outcomes.

A literature review and position-point paper (Narici et al. 2020) summarizes impacts of sedentarism on neuromuscular, cardiovascular and metabolic health due to the COVID-19 home confinement. Regular PA is very important, because the opposite behavior, sedentarism, leads to a rapidly loss of muscle mass, degenerative changes of neuromuscular system, a reduced cardiorespiratory fitness and an increased rate of mortality. Practical recommendations in the phase of the COVID-19 pandemic are high volume resistance exercise with low to medium intensity, walking more than 5.000 steps per day (with an advised option to start monitor daily activity with technical devices, e.g. smartwatch/phone) and conduct outside PA as often as possible (e.g. walking).

The work of Pecanha et al. (2020) describes the social isolation and the increase in physically inactivity during the COVID-19 pandemic, in the context of the global burden of cardiovascular disease. The assessment of smartwatch data reveals that the level of PA (steps per day) is reduced in the current pandemic, which accordingly increases cardiovascular risk factors. In the current situation it is not possible to be as active as recommended in the general guidelines (150 minutes moderate to mild PA or 75 min intensive PA per week or combination of both), because of reduced sports offers/ programs (closed gyms, parks, centers). The focus under these restrictions is on home-based PA programs, which should be supervised in vulnerable individuals (e.g. elderly, individuals with diseases). Additionally, wearable technologies (activity trackers through smartphones/ smartwatches) should be implemented, to provide an overview of daily activities. Research reveals that the usage is feasible and effective to reduce inactivity and ensure improvements in cardiovascular parameters in individuals with increased cardiovascular risk. It is important that individuals need to be educated and supported by new technological possibilities. Governmental actions need to be taken to promote the role of PA as a health-care priority in general, especially in times like these.

The longitudinal study from Pietrobelli et al. (2020) investigates the effects from the Covid-19 lockdown on lifestyle behaviors in children with obesity in Italy. Previous studies revealed, that the school environment reduces obesity risks, because it provides routine and structure in daily life. Consequently, summer vacations lead to a higher body mass index. The same phenomena was observed through the confinement period. The participants were less active and gained weight, because of an unhealthier lifestyle. Practical recommendations, such as telemedicine lifestyle programs and guidance from practitioners in medicine, were discussed to encourage families to implement or keep a healthy lifestyle in times of pandemics.

### Useful results which could be adopted for the present situation

To identify transferrable information for a pandemic restriction such as Covid-19, we created the following topics. The topic *Design of PA programs* should gather general ideas for the development of future programs in times of isolation, home-confinement and social distancing. Research reveals that programs to promote PA need to be adapted into the home environment. Therefore, the connection to the internet is a precondition, which should be established in all communities. The government task is to prioritise internet telecommunications, thus preventing isolation and improving wellbeing (Kearns et al., 2019). To overcome barriers of conducting PA, the benefits of exercise should be explained. Furthermore, realistic exercise goals should be established (Dunlap et al., 1999). PA interventions for the elderly should be encouraged the afternoon, because at this time most elderly have a sedentary behavior (Leask et al., 2015).

The *examples of non-digital exercises and PA programs* demonstrate currently available programs, which could be adapted into the conditions during the time of a pandemic. One example is an indoor gardening program, because this activity can improve the life satisfaction, social networking and reduce the perception of loneliness (Mimi Mun Yee, 2010). Another possibility is Tai Chi, because it can be adapted to a wide variety of age skill levels, and be modified for seated exercise or performed standing with or without walker support. Furthermore, the exercises can be performed individually or in a group (Hallisy & Hallisy, 2018).

The *digital health technologies* focus on technical solutions, which could be implemented one-to-one for the isolation, home-confinement and social distancing. A possible home-based dual task training health program, principally for elderly, is “SocialBike”. The users cycle on a stationary bike, while they recognize objects/ animals along the road. An important feature is the possibility to train with others, to reduce the risk of social isolation (Brady et al., 2020). Exergames demonstrate beneficial effects for individuals with serious mental illnesses, for promoting physical health, improving self-efficacy and an increase in social integration (Dobbins et al., 2020).

The topic *Space-Simulation Isolation* is a scenario which can be compared to the current pandemic. The research in this field shows that sedentary behavior and bed confinement should be overcome with performing exercises, such as resistance training, for the prevention of psycho-physiological changes (Goswami, 2017; Schneider, 2010).

*Real-Life Isolation*, describes situations in which individuals experience the confinement-situation not only in a pandemic, also in real-life. For example, homeless individuals are often isolated from the society. But their isolated experience can be conquered with the performance of activities, which lead to social interaction and better general health behavior (Malden et al., 2019). Another example is inmates. Results indicate that PA increases the level of self-control of a person in prison. Higher levels of exercise are connected to less psychological stress among older inmates. This knowledge is important for the characteristics of a pandemic.

## Discussion

The primary goal of this review was to identify practical recommendations of studies for “Staying Active” during Covid-19. We can summarize that regular daily routines and exercises should be carried out in daylight. When it is allowed, people should perform their exercises as often as possible outside, in nature (e.g. walking or jogging). For monitoring the steps per day (5,000 steps per day should be a realistic goal for healthy individuals), activity trackers should be used. The intensity of all activities should be mild to moderate. Other exercises, such as strength training, should be in addition, conducted with an intensity from low to medium. Further, relaxion activities are relevant to reduce the stress-level, which is in times of this pandemic often high (Rajkumar, 2020).

Our search indicates, that community-based programs have the potential to decrease the feeling of isolation and the stress-levels. In the situation of home-confinement, exergames, are a perfect innovation which should have the integrated function of a group play modus. Effects are higher levels of PA and an increase of a feeling of social belonging for older adults, including those with mental illness. Smart technologies also have the potential to better manage and understand health conditions. It should be considered that the family doctor have access to the results of the PA program and the health condition, such as results from a heartrate monitor. With, or even without internet-assess, Tai Chi is another possible exercise which can be adapted to different skills and different ages. Dancing shows positive effects, such as a reduction of the feeling of social isolation, especially for patients with depressions. Telephone supervision or text messages should be established between health-care professionals and patients. Exercises are useful to counteract against psycho-physiological changes and higher self-control -level during phases of confinement.

In relation to the secondary question, we generated 5 topics which bundle information from examples of non-digital and digital PA programs, space-simulation, and real-life isolation scenarios, which all present useful results adoptable to the pandemic restrictive conditions. When creating a PA program, it is important to explain first about the benefits of the exercise. Realistic individual goals need to be established. Notably digital community exercise programs have the potential to motivate individuals and to reduce their feeling of loneliness, even if they train from their home. Best scenarios would integrate family members, because this leads to more motivation and an increased effort time for conducting PA. Government actions should promote PA as a health-care priority. Especially to vulnerable individuals, such as elderly and patients with diseases who may require a longer duration of isolation, in order to avoid virus transmission.

For phases of home-isolation in the pandemic, practical materials for possible exercises of home-based programs are recommended, which can be accessed from the World Health Organisation (WHO), the American Heart Association (AHA), or the American College of Sports Medicine (ACSM): The training advice from the WHO (2020c) are general (e.g. exercise classes online; dance to music; play active video games; strength & balance training). For duration, they advise for health adults 30 minutes per day PA, and for children one hour. In addition to that, some practical ideas that should be established are communicated, such as regular standing breaks, walking up and down the stairs, or the use of skipping ropes. The AHA invites people to join on virtual workshop (AHA, 2020a) for different workouts (e.g. full-body workout, lower body tone workout, stretch workout). In addition, they offer useful exercises and explanations in their videos, such as a sofa stretch or chair dips. Moreover, they compare activities in and around the house for times of pandemic restrictions with the following real-life-scenarios:

- “10 minutes of stretching is like walking the length of a football field”
- “2.5 hours of walking every week for a year is like walking across the state of Wyoming”
- “30 minutes of singles tennis is like walking a 5K”
- “1 hour of dancing every week for a year is like walking from Chicago to Indianapolis”
- “20 minutes of vacuuming is like walking one mile”
- “30 minutes of grocery shopping every other week for a year is like walking a marathon” (AHA, 2020b)

Through these comparisons, the reader can be motivated to be more active, which is an important pre-condition for conducting PA. They have a home-based exercise program (AHA, 2020c), with one challenge targeted at children to be active at home (AHA, 2020d). Practical tips for the whole family are discussed, such as active chore cards, fitness during TV, active games (e.g. twister), or playing with pets (AHA, 2020e).

The ACSM lists at the beginning of the page some links to other blogs and videos for staying active at home during Covid-19 (ACSM, 2020a). The rest of the content is rather general about recommended activities in a normal situation, omitting the pandemic restrictions.

## Conclusion

The reduced PA levels, along with increased sedentary behavior have been identified as a crucial consequence of home confinement in pandemic time to overall health. This review aimed to summarise the literature for practical recommendations on staying active during times of a pandemic. Some effort has been made to help people find some alternatives to ensure that they can stay active during such times. The results provided in this review demonstrate a small but rapidly growing body of evidence around some practical recommendations for performing PA in times of pandemics such as Covid-19. The information, especially those from WHO and AHA are for the current pandemic really helpful. When comparing the results from the different organisations and the reviewed literature, there is a gap which need to be filled with special recommendations for older adults and individuals with diseases, with explanations on the task, duration, intensity and positive effects of the exercises. This knowledge needs to be disseminated more broadly into the world. Also, to be considered, are the development of exergames which combine a social belonging element with PA programs at home in times of home confinement and isolation. This paper helps researchers, to identify activities to recommend in times of home-confinement.

## Data Availability

Availability of all data referred to in the manuscript is ensured.

